# Longitudinal Cognitive Recovery in Survivors of Critical Illness: Impact of Sepsis and Benzodiazepine Exposure

**DOI:** 10.1101/2025.09.23.25336354

**Authors:** Ruhi Sahu, Ruth-Ann Brown, Anthony S Bonavia

## Abstract

**Background:** Post-critical illness cognitive dysfunction (PCICD) is a common and debilitating condition affecting survivors of critical illness. While sepsis has been implicated in poor cognitive outcomes, its independent contribution remains unclear due to multiple associated confounders in critical illness. This study aimed to characterize cognitive recovery trajectories over 12 months post-intensive care unit (ICU) and to evaluate the influence of sepsis and benzodiazepine exposure on cognitive outcomes.

**Methods:** In this single-center, prospective cohort study, adult ICU survivors were assessed at 30 days, 3 months, 6 months, and 12 months post-discharge using the telephone-administered Mini-Mental State Examination (MMSE) or Montreal Cognitive Assessment (MoCA). Scores were standardized into *z-*scores for comparability. Mixed-effects models assessed changes over time and the effects of clinical covariates, including sepsis status and benzodiazepine exposure. Additionally, we investigated whether any one specific cognitive domain was disproportionally impaired by critical illness over time.

**Results:** Of 197 eligible patients during the enrollment period, 77 (39%) completed at least one cognitive assessment. Standardized cognitive scores significantly improved over time, with the greatest gains observed within 6 months: +0.40 SD at 3 months (p = 0.041), +0.54 SD at 6 months (p = 0.016), and +0.49 SD at 12 months (p = 0.033) compared to scores at the time of acute illness. Sepsis status had no significant effect on recovery trajectory. No single cognitive domain was disproportionately affected by critical illness; instead, changes were observed in the overall score over time. Benzodiazepine exposure showed complex associations: longer duration (–0.24 SD/day, p = 0.008) and higher daily dose (–0.02 SD/unit, p = 0.006) were linked to worse cognition, while total cumulative dose was paradoxically associated with better scores (+0.03 SD/unit, p < 0.001), possibly reflecting confounding by indication or survival bias.

**Conclusions:** ICU survivors experience gradual cognitive recovery over the first year, primarily within 6 months. Sepsis does not independently affect this trajectory. Benzodiazepine exposure, especially prolonged or high daily dosing, emerges as a modifiable risk factor for cognitive impairment, consistent with prior investigations of PCICD. These findings highlight the importance of sedation strategies and structured cognitive follow-up.

## Introduction

As survival after critical illness improves, long-term cognitive sequelae have become a major public health concern. Post-critical illness cognitive dysfunction (PCICD) encompasses persistent deficits in attention, memory, executive function, and processing speed that may last for months to years following discharge from the intensive care unit (ICU). These impairments are both common and disabling, limiting quality of life, independence, and return to work. Cognitive impairment after ICU admission is also taxing to patients and their families and carries an enormous societal cost estimated at $18 billion per year^1^. Risk is heightened in ICU-related conditions such as sepsis, shock, and respiratory failure, and is compounded by exposures including prolonged mechanical ventilation, sedative and steroid use, immobility, and comorbid disease^2,3^.

Meta-analyses estimate that nearly 50% of patients experience cognitive impairment within one month of ICU discharge, with 28.3% still impaired more than one year later. Honarmand et al. similarly reported PCICD in ∼46% of survivors two years after discharge^4^. These findings underscore the need to map cognitive recovery trajectories, particularly in the first 3–6 months post-discharge when interventions may be most effective.

Sepsis, one of the most common ICU admission diagnoses, has been implicated in long-term PCICD. Sepsis survivors may exhibit higher rates of cognitive dysfunction than non-septic patients, with one study reporting up to a three-fold increased risk^5^. Conversely, a multicenter cohort of mechanically ventilated patients found no significant difference in cognitive, psychiatric, or quality-of-life outcomes at six months between septic and non-septic survivors of similar illness acuity^6^. Thus, the independent effects of sepsis on cognitive decline remain uncertain.

Other clinical factors—including age, comorbidity burden, delirium, and educational level—also shape cognitive outcomes. Advanced age and higher Charlson Comorbidity Index scores increase risk, whereas higher education appears protective; patients with more than 12 years of education had up to a 95% lower odds of PCICD compared with those with six or fewer years^7^. Delirium is the most consistent modifiable predictor, independently associated with long-term decline, mortality, and prolonged ICU stay, with an estimated patient-level 30-day cumulative cost of $18,000^8-10^. Benzodiazepine exposure, which increases delirium risk, has been identified as another potentially modifiable contributor ^8^.

Despite growing evidence, important gaps remain. The longitudinal course of specific cognitive domains within global PCICD—particularly in septic versus non-septic patients—remains poorly characterized, and the influence of benzodiazepine exposure is not well understood. To address these gaps, we conducted a prospective cohort study of ICU survivors, assessed at 30 days, 3 months, 6 months, and 12 months post-discharge using validated cognitive tools. We examined recovery trajectories, stratified by sepsis status and benzodiazepine exposure, to identify predictors of persistent or progressive impairment.

## Methods

### Study Design and Setting

A single-center, prospective cohort study was conducted in the medical and surgical intensive care units (ICUs) of an academic medical center from September 2020 to August 2024. Consecutive adult patients (≥ 18 y) who survived an index episode of critical illness and were discharged alive from the ICU were screened. Sepsis and septic shock were defined by Sepsis-3 criteria^11,12^. Exclusion criteria were: (1) pre-morbid severe neuro-cognitive disorder prohibiting assessment, (2) non-English speakers, and (3) refractory delirium precluding follow-up.

### Cognitive Assessments

Cognitive function was evaluated via telephone interviews at 30 days and again at 3, 6, and 12 months after ICU discharge using two standardized instruments. Initially, we administered the Mini-Mental State Examination (MMSE; score range 0–22; Appendix A). In September 2022, we transitioned to the blind/telephone Montreal Cognitive Assessment (MoCA v8.1 Blind/Telephone^13^; score range 0–27). The transition from MMSE to MoCA was motivated by methodological advantages inherent to the MoCA instrument. Unlike the MMSE, the MoCA encompasses more complex cognitive tasks, including an abstraction component, making it more sensitive for detecting subtle cognitive deficits frequently observed among ICU survivors. Additionally, the MoCA version used in this study was specifically designed to facilitate telephone administration, allowing for consistent, remote follow-up assessments. Both tools assessed orientation, attention, delayed recall, and language; the MMSE additionally included an immediate recall task, whereas the MoCA incorporated an abstraction component. The MoCA assessment was administered by a trained/certified research coordinator.

### Benzodiazepine Exposure

Given the observational nature of this study, benzodiazepine exposure was converted to lorazepam equivalents (LE) using route-specific factors: intravenous (IV) midazolam = 0.5x, oral (PO) midazolam = 0.1x, lorazepam (IV/PO) = 1x. The IV factor reflects RCT-based equipotency (lorazepam ≈2x midazolam) and published ICU conversion tables^14^; the PO factor conservatively accounts for midazolam’s low, variable oral bioavailability and relatively stable oral lorazepam bioavailability, with additional variability in enteral absorption during critical illness^15-17^. To assess robustness to alternative published ratios, we performed a sensitivity analysis using IV midazolam = 0.33x^18,19^.

### Statistical Analysis

All analyses were conducted using Python v3.11, *pandas* v2.2, *statsmodels* v0.14, and *scikit-learn* v1.5 libraries. Raw scores from the MMSE and MoCA assessments were standardized into *z-*scores using instrument-specific normative means and standard deviations, placing both instruments on a common metric and facilitating direct comparisons. *Z-*scores were calculated as follows:

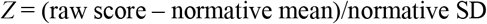

All subsequent analyses were performed on these standardized values. Missing data were managed using listwise deletion, whereby observations with missing cognitive assessment scores at any given time point were excluded from analyses. Specifically, when constructing the longitudinal dataset for both total and domain-specific cognitive scores, subjects without available assessments at a particular visit were omitted from that visit’s analysis but retained for other visits for which data were available. Mixed-effects models inherently accommodate unbalanced data and partially observed trajectories, mitigating potential bias from intermittent missingness under the assumption that missing data were missing at random.

To characterize the unadjusted cognitive recovery trajectory, a linear mixed-effects model with restricted maximum likelihood estimation was implemented. Standardized cognitive *z-*scores served as the dependent variable, modeled as a function of assessment time points (categorical: 30 days, 3 months, 6 months, 12 months), cognitive test type (MoCA vs. MMSE), and their interaction term. Subject-specific random intercepts and visit-specific random slopes accounted for the correlated nature of repeated assessments. Estimated marginal means at each follow-up time point were contrasted with the 30-day baseline.

The probability of cognitive impairment at 12 months was explored using penalized logistic regression, defining impairment as MoCA <26 or MMSE <24. This model quantified the odds of impairment per standard deviation increase in *z-*score, reported as an odds ratio.

To examine potential differences in recovery trajectories between septic and non-septic patients, we conducted a sepsis-stratified mixed-effects analysis, substituting cognitive test type with sepsis status (yes/no) and incorporating a visit-by-sepsis interaction.

Finally, a fully adjusted mixed-effects model was constructed, including *a priori* selected clinical covariates: age, Charlson Comorbidity Index, peak SOFA score, ICU length of stay, benzodiazepine administration, sepsis status, cognitive assessment tool, and their interactions. Results were reported as fixed-effect estimates accompanied by 95% confidence intervals. Statistical significance was defined as a two-sided *P-*value <0.05. Model assumptions were verified by inspecting residual quantile–quantile plots, with no significant deviations noted. Visualizations, including line plots with 95% CIs, boxplots, waterfall plots, and forest plots, were generated using *seaborn* v*0*.*13*.

## Results

### Cohort and Assessments

Of 197 eligible survivors, 77 (39%) completed at least one cognitive assessment and were included in the analysis (**Table 1**). These assessments yielded a longitudinal dataset with a mean of 2.6 cognitive assessments per subject. After limiting analysis to cases having all clinical covariates, the mixed□effects model was fit on 65 subjects.

**Table 1.** Clinical Features of Critically Ill Patients With and Without Sepsis.

|  | **Sepsis cohort**<br>**(n = 48)** | **Non-sepsis cohort**<br>**(n = 29)** |
| --- | --- | --- |
| Age (mean, IQR) | 66.1 (57.5, 77.0) | 65.2 (59.0, 75.5) |
| Charlson Comorbidity Index (mean, IQR) | 4.4 (2.0, 7.0) | 4.0 (2.0, 6.0) |
| SOFA score (mean, IQR) | 6.9 (4.0, 9.8) | 7.5 (5.5, 10.0) |
| ICU length of stay (mean, IQR) | 3.5 (1.0, 4.0) | 6.4 (2.0, 8.5) |
| Delirium, by CAM-ICU, during ICU stay (n, %) | 5.0 (10.4) | 6.0 (20.7) |
| Number of days intubated in the ICU (mean, IQR) | 0.9 (0.0, 0.0) | 2.0 (0.0, 1.0) |
| Days of GCS<15 during ICU stay (mean, IQR) | 0.8 (0.0, 1.0) | 2.2 (0.0, 3.0) |
| Nadir GCS score (mean, IQR) | 14.3 (14.0, 15.0) | 13.0 (13.0, 15.0) |
| Patients receiving benzodiazepines during ICU (n, %) | 9.0 (18.8) | 10.0 (34.5) |
| Mean number of days of benzodiazepine use, *among patients receiving benzodiazepines* (mean, IQR) | 2.0 (1.0, 2.5) | 2.5 (1.0, 3.8) |
| Highest education level (n, %) |  |  |
| ---high school or less | 8.0 (16.7) | 5.0 (17.2) |
| ---college | 6.0 (12.5) | 11.0 (37.9) |
| ---post-graduate | 2.0 (4.2) | 1.0 (3.4) |
| ---unknown | 32.0 (66.7) | 12.0 (41.4) |
| Patients having baseline cognitive deficit (n, %) | 6.0 (12.5) | 3.0 (10.3) |
| Patients deceased <90 days (n, %) | 1.0 (2.1) | 0.0 (0.0) |
| Patients with a cognitive assessment at 30 days (n, %) | 31.0 (64.6) | 19.0 (65.5) |
| Patients with a cognitive assessment at 3 months (n, %) | 34.0 (70.8) | 22.0 (75.9) |
| Patients with a cognitive assessment at 6 months (n, %) | 31.0 (64.6) | 14.0 (48.3) |
| Patients with a cognitive assessment at 12 months (n, %) | 27.0 (56.2) | 16.0 (55.2) |

### Overall Recovery Trajectory

Standardized *z-*scores rose significantly from 30 days to: 3 months by +0.40 SD (95 % CI 0.015–0.779; p=0.042), 6 months by +0.54 SD (95 % CI 0.097–0.982; p=0.017), and 12 months by +0.49 SD (95 % CI 0.028– 0.951; p=0.038) (**Fig 1A**). There were no significant effects of test type (MoCA vs. MMSE; β = 0.009 SD, p = 0.97), and no significant sepsis-time interactions (all p > 0.20), indicating comparable trajectories across instruments and sepsis status (**Figs 1A–B, 4**). Spaghetti plots (**Fig 1C**) and box plots (**Fig 1D**) corroborated these improvements.

**Fig 1.**
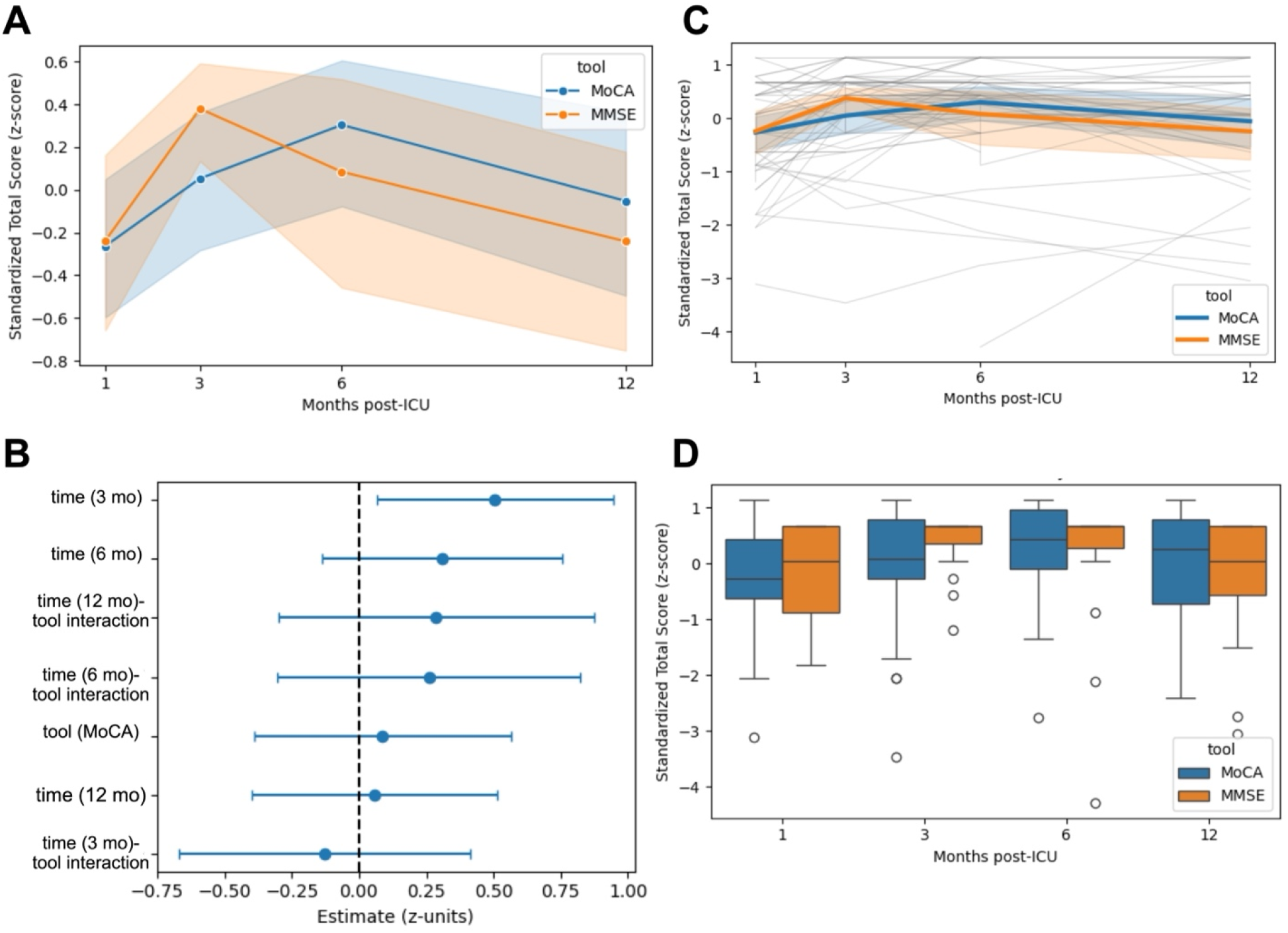
Recovery of Cognitive Scores, as measured by MoCA and MMSE. (A) Standardized Total Score recovery, showing mean *z-*scores at 1, 3, 6 and 12 mo post-ICU, with 95% CI from a categorical time mixed effects-model. No model assumptions other than independence. (B) Forest plot for fixed effect coefficients. Point estimates and 95% CIs for fixed effects from the continuous mixed□effects model (time, tool, time-tool interactions). The vertical line at zero denotes no effect. (C) Spaghetti pot of individual patient cognitive trajectories. Gray lines for each patient’s *z-*score trajectory and overlaid colored mean trajectories ±95% CI. (D) Boxplot distribution over time of *z-*scores at each time point (IQR ± 1.5 times IQR, with outliers). n (MoCA/MMSE): 1 mo = 34/16; 3 mo = 38/19; 6 mo = 27/19; 12 mo = 24/19.

**Fig 2.**
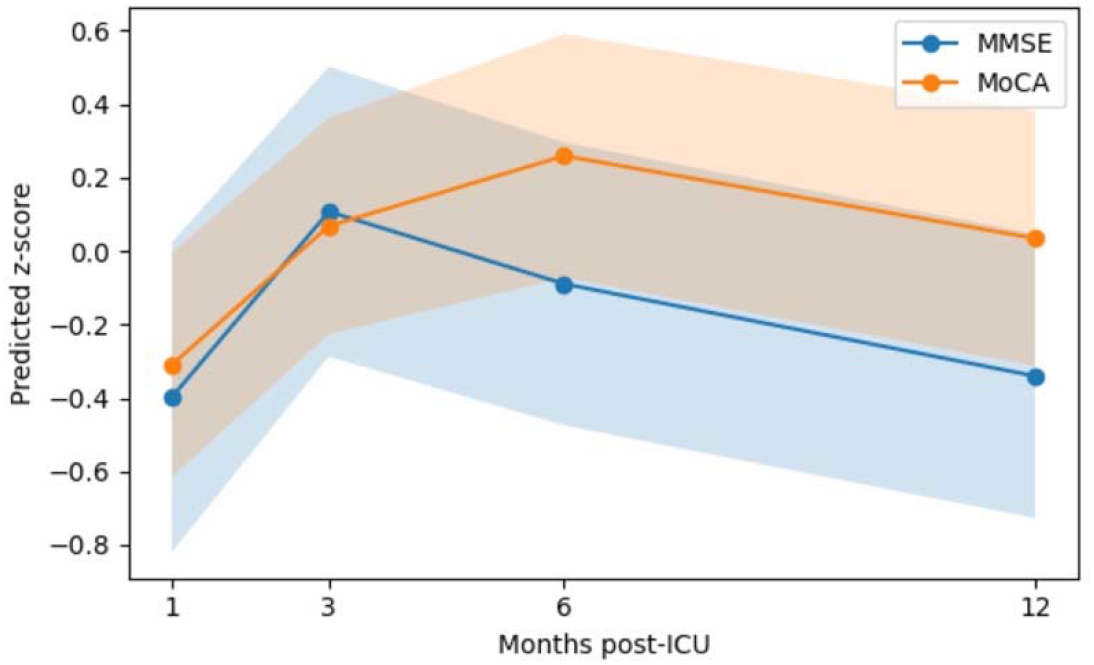
Estimated Marginal Mean Recovery Over Time. Model-predicted *z-*score trajectories at 30 days, 3 months, 6 months, and 12 months— derived from a linear mixed effects model that time visit as a continuous variable. Solid lines represent the estimated marginal means (EMMs) of standardized cognitive scores per month; shaded ribbons denote 95 % confidence intervals. Unlike Fig 1, which plots observed, mean *z-*scores at each discrete time point, this panel illustrates the smoothed, model-based trend in cognitive recovery across the entire 12-month period.

### Adjusted Clinical Predictors

Cognitive performance, by standardized *z-*scores, improved over time among ICU survivors, with significant gains observed at 3-, 6-, and 12-months post-discharge compared to the 1-month assessment (**Table 2**). Greater benzodiazepine exposure during critical illness was consistently associated with worse cognition: each additional day of use was linked to a –0.24 SD decline (p = 0.008), and higher average daily dose was similarly detrimental (–0.02 SD/unit, p = 0.006). Interestingly, total cumulative benzodiazepine dose was positively associated with cognitive scores (+0.03 SD/unit, p < 0.001), possibly reflecting confounding by indication or survival bias. Other covariates—including age, comorbidity burden, illness severity, duration of impaired consciousness, baseline cognitive status, and assessment tool used—were not significantly associated with outcomes (all p > 0.20).

**Table 2.**
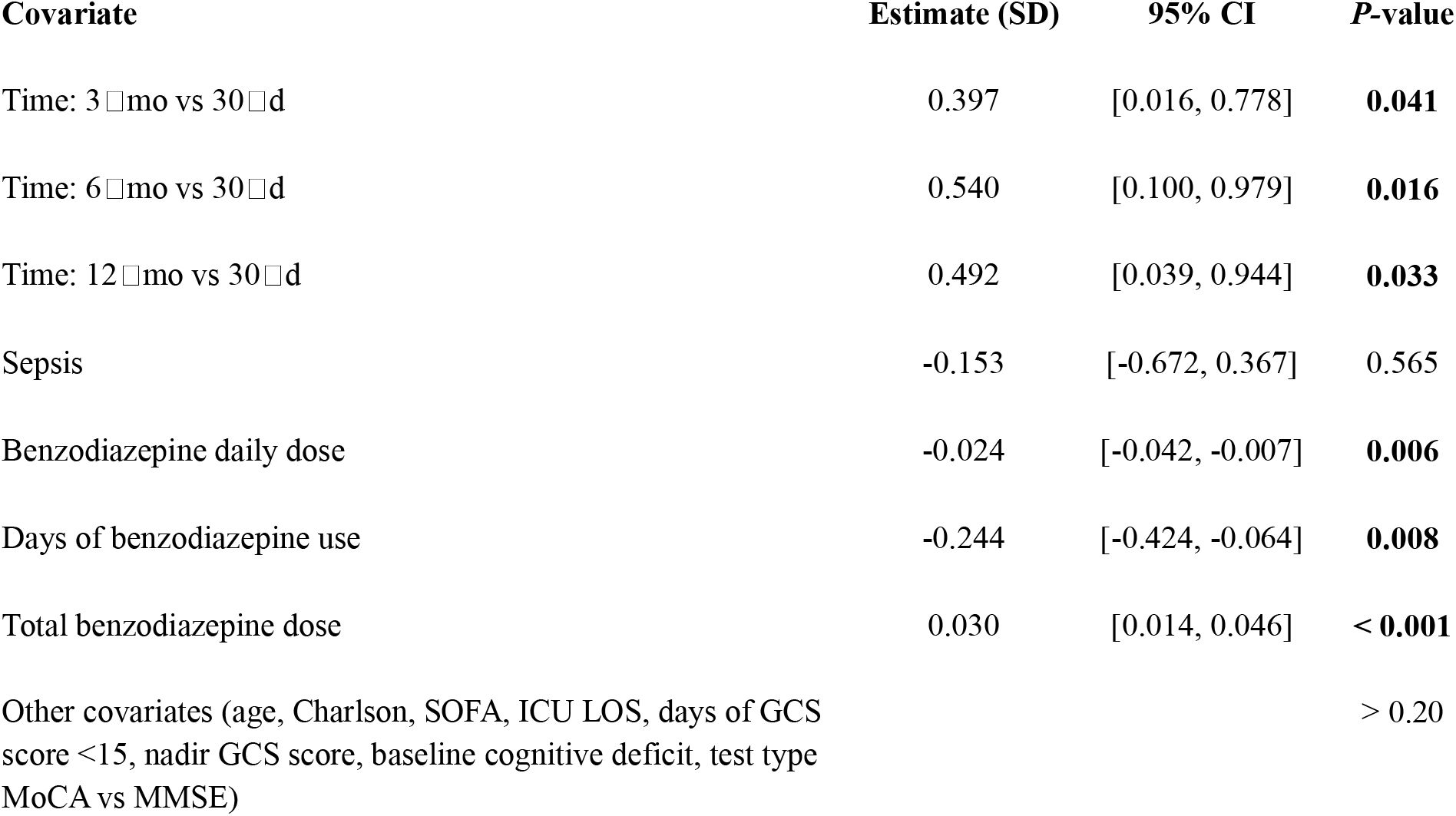
Adjusted Associations Between Clinical Covariates and Longitudinal Cognitive Outcomes. Estimates reflect standardized cognitive performance (*z-*scores) in a mixed-effects model with random intercepts. Positive estimates indicate improved cognitive scores.

Sepsis status had no significant main or interaction effects (baseline β = –0.153 SD, p = 0.56; sepsis-time interactions all p > 0.20), confirming similar recovery trajectories between septic and non-septic patients (**Fig 3**).

**Fig 3.**
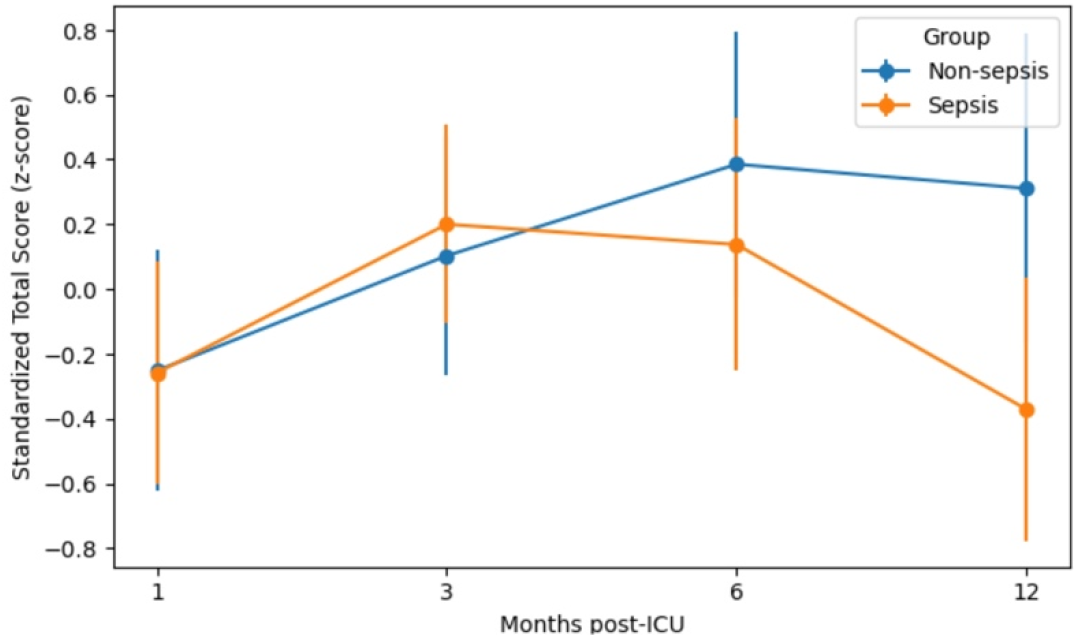
Cognitive Recovery in Sepsis vs Non-Sepsis patients. Mean *z-*scores ±95% CI at 1, 3, 6 and 12 months. n (non-sepsis/sepsis): 1 mo = 19/32; 3 mo = 22/35; 6 mo = 14/33; 12 mo = 15/29.

### Domain□Specific Trajectories

When modeled separately, none of the six cognitive domains (Language, Attention, Abstraction, Orientation, Immediate Recall, Delayed Recall) demonstrated a statistically significant change in standardized scores over the first year (**Fig 4**). Abstraction showed the largest positive slope (β = 0.039 *z*-units per month, SE = 0.023, p = 0.087), and Orientation trended upward (β = 0.024, SE = 0.013, p = 0.065), but neither reached conventional significance. Attention (β = 0.008, p = 0.54), Immediate Recall (β = –0.005, p = 0.86), Delayed Recall (β = –0.004, p = 0.79), and Language (β = –0.008, p = 0.58) remained essentially flat. Models used subject□level random intercepts (mixed□effects) when identifiable, with ordinary least squares fallback if convergence failed.

**Fig 4.**
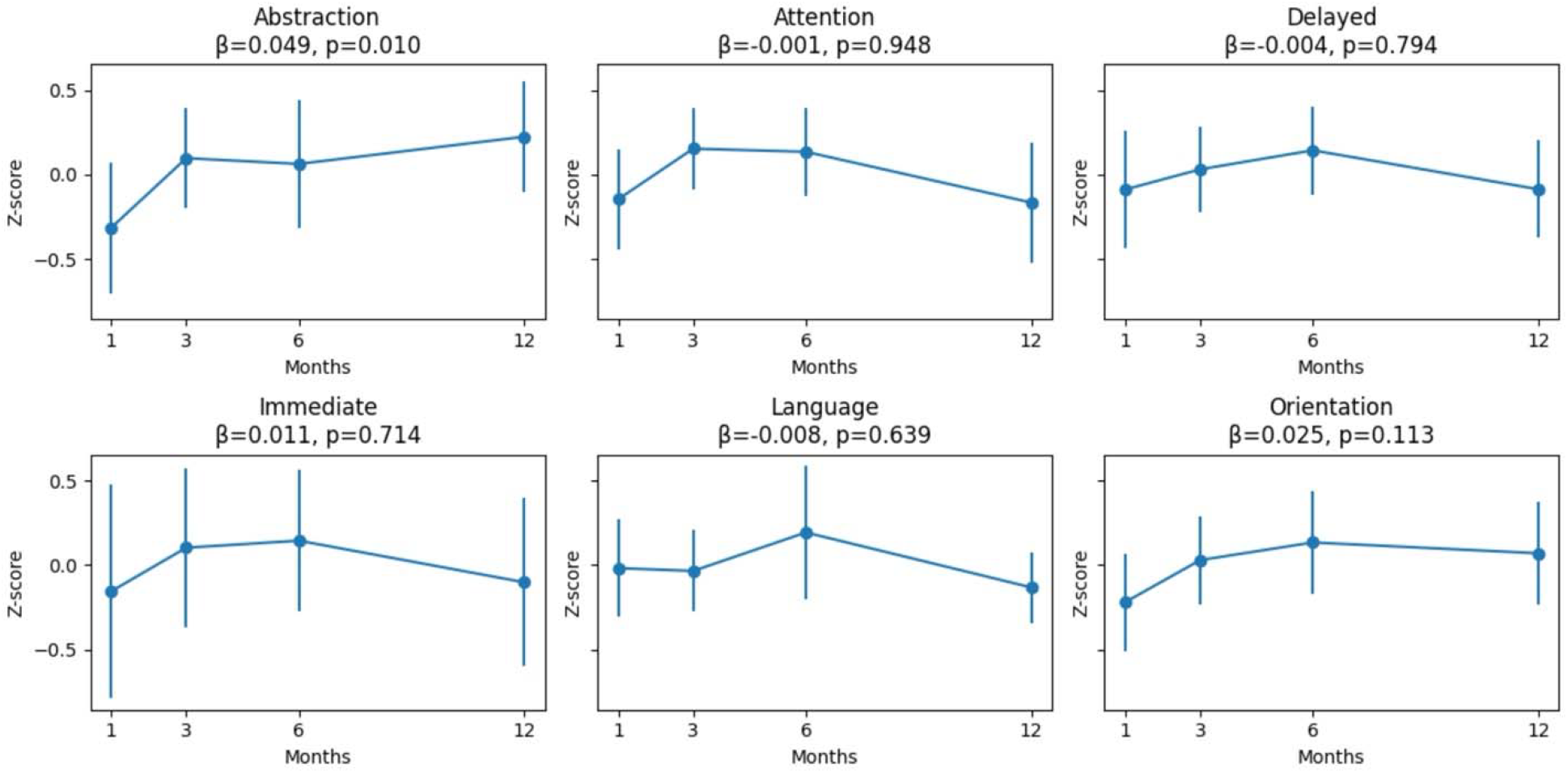
Small□Multiples of Cognitive Domain Recovery. Each panel shows the mean standardized (*z-*scored) performance in one cognitive domain (Abstraction, Attention, Delayed Recall, Immediate Recall, Language, Orientation) at 30 d, 3 mo, 6 mo, and 12 mo post□ICU. Points indicate the mean; vertical bars denote 95 % confidence intervals (mean ± 1.96 x SE). In each subplot title, the estimated slope (β) and *P-*value for the time effect is shown, derived from a linear mixed□effects model with a random intercept (or, where the mixed model did not converge, from ordinary least squares). No domain exhibited a statistically significant improvement over the study period. Immediate = Immediate Recall; Delayed = Delayed Recall.

## Discussion

This prospective longitudinal cohort study provides additional insights into cognitive recovery among ICU survivors. Our primary finding was that cognitive performance improved significantly over the first year after ICU discharge, with most recovery occurring within the first six months. These findings are consistent with prior studies ^20^. The impairment was not confined to any single domain but reflected a global pattern of dysfunction. Sepsis status did not significantly impact the trajectory or magnitude of recovery. Recent research similarly suggests that sepsis alone may not be a strong independent predictor of long-term cognitive impairment after critical illness ^6,21^.

Benzodiazepine exposure presented a complex picture. Longer duration and higher average daily dose of benzodiazepines were linked to worse cognitive outcomes. This aligns with extensive literature showing benzodiazepines increase the risk of delirium and subsequent cognitive decline ^22-24^. Surprisingly, total cumulative benzodiazepine dose was positively associated with better cognitive scores. This unexpected finding may reflect confounding by indication or survival bias: patients who were alert or stable enough to receive higher cumulative doses may have been inherently more likely to survive and participate in follow-up assessments. Alternatively, benzodiazepines may have contributed to patient comfort, reduced ICU-related stress, or prevented agitation-related complications that indirectly supported cognitive recovery in select individuals.

This study has several limitations. First, it was conducted at a single academic ICU with a modest sample size which may limit generalizability. The cohort may not reflect the diversity of case-mixes or rehabilitation practices seen in community or non-academic settings. The study is under-powered to detect subtle differences, particularly in interaction terms across subgroups such as sepsis vs non-sepsis patients.

Second, participant attrition and unbalanced follow-up introduce the potential for bias. While we employed linear mixed models that can handle data under the “missing at random” assumption, only 65% of participants completed the 30-day assessment and 56% remained at the 12-month timepoint. If sicker patients were less likely to return for testing, our estimates may understate the true cognitive burden. Moreover, data missingness may have been non-random: participants with greater cognitive impairment were likely underrepresented because they were less likely to answer or complete telephone assessments.

Third, we lacked formal cognitive baseline assessments prior to ICU admission. Although we standardized scores within each cognitive tool, we cannot determine whether individuals recovered to their premorbid cognitive level. This absence limits interpretation of whether changes over time reflect true cognitive decline or return to baseline function.

Fourth, we were unable to adjust for all relevant covariates due to missing data. Initial models attempted to include clinical predictors such as sepsis status, age, Charlson comorbidity index, peak SOFA score, days of benzodiazepine exposure, ICU length of stay, and education level. Charlson comorbidity index had < 1 % missingness; however, high rates of missing data for education level (53 % missing) reduced usable observations. To retain power, the final longitudinal model was pared down to fixed effects for time (visit), tool and their interaction, plus a random intercept for subject. Consequently, residual confounding by illness severity, sedation exposure, delirium duration or socio-economic factors cannot be excluded.

Fifth, we transitioned from MMSE to MoCA and applied equipercentile linking to harmonize scores. While this approach has been validated in general populations^25-29^, our linking curves were derived from the same study cohort and not externally validated Additionally, misclassification may have occurred due to the conventional impairment cut-offs (<26 for MoCA, <24 for MMSE), which may have suboptimal sensitivity and specificity in post-ICU populations.

Despite these limitations, our study adds to the growing literature on cognitive trajectories following critical illness, offering a unique perspective on the potential role of benzodiazepines. Future research with larger, multicenter cohorts and more comprehensive covariate data is warranted to further explore these associations.

## Data Availability

All data produced in the present work are contained in the manuscript

## Acknowledgements

Not applicable.

## Author Contributions

RS and ASB conceived the study. ASB designed the study. RB and RS curated the data, while RS and ASB performed formal analyses. RS and ASB prepared the figures and drafted the manuscript. All authors reviewed and edited the final version. ASB supervised the project and secured funding.

## Statements and Declarations

### Ethical Considerations

This study was approved by the Penn State University Human Subjects Protection Office (IRB Protocol #15328, 7/30/2020). The studies were conducted in accordance with local legislation and institutional requirements. The study was also registered on clinicaltrials.gov (NCT03146546).

### Consent to Participate

All participants, or their legally authorized representatives (LARs), provided informed consent prior to study enrollment. Written consent was obtained whenever feasible. In situations where the LAR could not be physically present and timely enrollment was required, verbal consent was obtained by the study coordinator in accordance with IRB approval.

### Consent for Publication

Not applicable.

## Appendices

### Appendix A: The Mini-Mental Status Examination (MMSE)

1. What is the year/season/date/day/month (score 0-5, *Orientation* Domain)
2. Where are you (country/State/Town/hospital or bldg/floor or street (score 0-5, *Orientation* Domain)
3. I am going to name 3 things and I want you to remember them. Fish, hook, shoe, green. First, can you repeat these? (score 0-3, *Memory - Immediate Recall* Domain)
4. Can you count backward from 100 by 7 (stop after 5 answers, score 0-5, *Attention* Domain)
5. Can you spell “world” (as in, the world we live in) backward? (score 0-5, one point per letter, *Attention* Domain)
6. Can you remember the 3 things I had asked you to remember before (score 0-3, *Memory - Delayed Recall* Domain)
7. Can you repeat the following: “no if, and or buts” (score 0-1, *Language* Domain) *Total Mini-Mental Status Exam Score (range 0-27)*

